# Alzheimer’s Disease Heterogeneity Explained by Polygenic Risk Scores Derived from Brain Transcriptomic Profiles

**DOI:** 10.1101/2022.12.20.22283737

**Authors:** Jaeyoon Chung, Nathan Sahelijo, Toru Maruyama, Junming Hu, Rebecca Panitch, Weiming Xia, Jesse Mez, Thor D. Stein, the Alzheimer’s Disease Neuroimaging Initiative, Andrew J. Saykin, Haruko Takeyama, Lindsay A. Farrer, Paul K. Crane, Kwangsik Nho, Gyungah R. Jun

## Abstract

**INTRODUCTION:** Alzheimer’s disease (AD) is heterogeneous, both clinically and neuropathologically. We investigated whether polygenic risk scores (PRSs) integrated with transcriptome profiles from AD brains can explain AD clinical heterogeneity.

**METHODS:** We conducted co-expression analysis and identified gene-sets (modules) which were preserved in three AD transcriptome datasets and associated with AD-related neuropathological traits for neuritic plaques (NPs) or neurofibrillary tangles (NFTs). We computed the module-based PRS (mbPRS) for each module and tested associations for mbPRSs with cognitive test scores, cognitively-defined AD subgroups, and brain imaging data.

**RESULTS:** Of the modules significantly associated with NPs and/or NFTs, the mbPRSs from two modules (M6 and M9) showed distinct associations with language and visuospatial functioning as well as their matching AD-subgroups and brain atrophy at specific regions.

**DISCUSSION:** Our findings demonstrate that polygenic profiling based on co-expressed gene-sets can explain heterogeneity in AD patients, enabling to genetically-informed patient stratification and precision medicine in AD.

## 1. BACKGROUND

Late onset Alzheimer disease (AD) is a complex disorder with clinical and neuropathological heterogeneity [1, 2]. Types of clinical heterogeneity include progression rate, predominant cognitive symptoms, and whether psychotic symptoms manifest [1]. AD neuropathology can also be varied with complications of other neuropathological traits beyond plaques and tangles [1, 2]. Clinical and neuropathological heterogeneity may have contributed to the repeated failure of AD clinical trials [3]. Classification of heterogeneous AD patients into biologically-relevant subgroups may improve our understanding of biological mechanisms underlying the variability of cognitive symptoms and trajectories of decline, as well as lead to development of subgroup-specific treatment options [4].

Different AD subtypes have been previously proposed based on neuropsychological and neuropathological characteristics [5-7], domain-specific cognitive functions, MRI brain imaging data [4], and metabolic profiling [8]. However, our understanding of molecular mechanisms underlying disease heterogeneity is still limited. Recent report illustrates that genetic variants with large effect sizes can distinguish six cognitively-defined subgroups of AD when compared with elderly controls [9]. A previous study showed that polygenic risk scores (PRSs) derived from clusters (*i*.*e*., gene-sets) in genome-wide association studies (GWASs) of type 2 diabetes (T2D)-related phenotypes have successfully classified T2D patients into different subtypes [10]. These studies demonstrate that PRSs from biologically connected gene-sets may explain disease heterogeneity and improve scientific understanding of biological mechanisms underlying disease subtypes. In addition, co-expression network analyses have shown to be useful for identifying biologically-connected and disease-relevant gene-sets using transcriptome data [11, 12]. Taken together, these findings led to the hypothesis that network analysis utilizing transcriptome data of AD brains could capture biologically relevant gene-sets responsible for distinct disease subtypes and PRSs derived from the gene-sets could explain clinical heterogeneity of AD.

In this study, we identified modules (sets of biologically relevant genes) by coexpression analysis and thereby generated module-based PRSs of AD patients. Then, using domain-specific cognitive functions, previously defined AD cognitive subgroups, and brain imaging data, we evaluated whether the module-based PRSs can explain cognitive impairment heterogeneity among the AD patients

## 2. METHODS

### 2.1. Sources of RNA sequencing data in autopsied AD brains for network analysis

Discovery coexpression analysis were performed using previously generated gene expression data from the dorsolateral prefrontal cortex (DLPFC) area of 65 autopsy-confirmed non-Hispanic white AD cases from the Framingham Heart Study and Boston University Alzheimer’s Disease Research Center (FHS/BUADRC) [13]. Details of procedures for quality control (QC) of RNA sequencing (RNA-Seq) data and neuropathological AD diagnosis are presented in **Supplementary Information** and previously reported elsewhere [13]. Additional RNA-Seq datasets for validation were obtained from the CommonMind portal (http://www.synapse.org) including post-QC normalized gene expression data (version #1) from the DLPFC area of 363 neuropathologically-confirmed AD cases in the Religious Orders Study and Rush Memory and Aging Project (ROSMAP) [14] and from temporal cortex area of 82 autopsy-confirmed AD cases in the Mayo Clinic Study of Aging (MAYO) [15].

### 2.2. Identifying Preserved and AD-associated Modules

Co-expression gene-sets (*i*.*e*., modules) were generated using the transcriptome data from the 65 AD brains in FHS/BUADRC using the Weighted Gene Co-expression Network Analysis (WGCNA) approach, which computes pairwise correlations for all gene pairs and clusters genes by the correlated expression levels [16]. Transcriptome data of AD-free controls were not included in our coexpression study because our interest is to identify gene-sets related with the disease heterogeneity, not the disease risk (*e*.*g*., cases versus controls). Details of co-expression module construction are presented in **Supplementary Information** and previously described [17]. Preservation of the discovery modules was evaluated in the two independent validation datasets, including ROSMAP and MAYO Clinic, using z-summary statistics [16]. We considered a module to be preserved if z-summary scores greater than 5.0 in both validation datasets [16]. Among the preserved modules, we selected AD-associated modules by enrichment analyses using gene-sets for AD phenotypes including AD-related neuropathological traits including neuritic plaques (NP) and neurofibrillary tangles (NFT) [18] and AD-risk [19]. We used AD associated genes for enrichment analyses that contained at least one single nucleotide polymorphism (SNP) with P<10^−3^ located within +/-20 kilobases from the gene for one of the AD phenotypes (NP, NFT, or AD-risk). Significant enrichment P-values<0.05 were applied using the Fisher’s exact test after false discovery rate (FDR) correction. Based on the result from the enrichment analysis for each module, we assigned the AD phenotypes (NP, NFT, or AD-risk) for which the module was most significantly enriched and used to calculate module based polygenic risk scores. The selected AD-associated modules were considered to generate module based polygenic risk scores.

We also examined expression coherence and cellular identifies of genes in each of the AD-associated modules using single cell RNA-seq data in five different cell types (astrocytes, microglia, oligodendrocytes, endothelia, and neurons) from the temporal lobe area (Gene Expression Omnibus ID: GSE67835) [20] and single nucleus RNA-seq data in seven cell types (astrocytes, microglia, oligodendrocytes, pericytes, endothelia, and excitatory/inhibitory neurons) from the prefrontal cortex in the ROSMAP [21]. Details of methods for deriving cell-type-specific gene-sets and their expression profiling are presented in **Supplementary Information** and reported elsewhere [13]. Enrichment of cell-type-specificity for each AD-associated module was tested using the Fisher’s exact test.

### 2.3. Genotypic and phenotypic data in ADNI

The Alzheimer’s Disease Neuroimaging Initiative (ADNI) is a longitudinal study assessing clinical, neuroimaging, genetic, and biomarker data from participants in various stages of cognitive impairment including cognitively normal (CN), mild cognitive impairment (MCI), and AD. Genetic and phenotypic data of ADNI participants were obtained from the LONI website (http://adni.loni.usc.edu). We used the ADNI genetic data for computing mbPRSs and phenotype data for evaluating the relationships between mbPRSs and cognitive impairment heterogeneity. Genome-wide genotype data from two different arrays (ADNI-1, n=679 and ADNI-GO/2, n=397) were imputed using the Haplotype Reference Consortium data. Details of quality control (QC), imputation, and population substructure procedures are described in the **Supplementary Information**. Characteristics of the sample after QC are presented in **Table S1**.

Since clinical spectrum of AD can be largely affected by impairment of specific cognitive functions [22], we hypothesized that deficits in particular cognitive domains explain at least in part disease heterogeneity (*i*.*e*., cognitive impairment heterogeneity). To explore this heterogeneity, we used domain-specific cognitive tests at the last exam from the ADNI dataset (**Table S1**): logical memory immediate (LIMMTOTAL) and delayed (LDELTOTAL) recall tests for *memory*; trail marking test A/B (TRAASCOR and TRABSCOR) for *executive functioning*; category fluency animal score (CATANIMSC) and Boston naming test total (BNTTOTAL) score for *language*; and clock test total score (COPYSCORE) for *visuospatial functioning*. Cognitive test scores were adjusted for age, sex, and education using linear regression, and the residuals derived from the regression models were ranked-transformed as previously described [17].

### 2.4. Computing and assessing polygenic risk scores for AD-associated modules in ADNI

We selected SNPs in each AD-associated module from the enrichment analysis for the assigned AD phenotype and generated module based polygenic risk scores (mbPRSs) using effect estimates of the selected SNPs for the assigned AD phenotype (NP, NFT, and AD-risk) from the enrichment analysis. For comparison, we also generated PRSs for three AD phenotypes (NP, NFT, and AD-risk) in a conventional approach, which aggregates effect estimates of SNPs with P<0.001 across the genome (*i*.*e*., genome-wide PRS [gwPRS]). Details about computing these two types of PRSs (gwPRS and mbPRSs) are included in the **Supplementary Information**.

After generating those PRSs in ADNI, we evaluated correlations among the mbPRSs and gwPRSs. To assess relevance of those PRSs to disease stages/progression, we stratified ADNI sample by disease stages (CN, MCI, and AD) at the last exam and compared mean values of PRSs between different disease stages. We also tested associations between PRSs and conversion status for disease progression (*e*.*g*., CN to MCI or AD; MCI to AD) excluding AD at baseline using logistic regression models after adjusting for age, sex, the first four PCs and the array information.

Next, we conducted association tests with mbPRSs or gwPRSs for specific cognitive domains using rank-transformed cognitive test scores as quantitative outcomes in linear regression models after adjusting the first four PCs and genotype platform as covariates. We followed up the nominally significant modules (P<0.05) with domain-specific cognitive test scores as cognitive impairment heterogeneity (CIH) modules.

We also attempted to replicate the associations between mbPRSs of the selected CIH modules and domain-specific cognitive test scores among 134 AD cases in FHS (dbGaP Study Accession ID: phs000056.v5.p3). Details of sample characteristics, imputation, computation of mbPRSs, and association tests with cognitive test scores in Neuropsychological Test Battery is described in the **Supplementary Information**.

### 2.5. Validating CIH modules with cognitively-defined AD subgroups in ADNI

Previously, 672 AD cases in ADNI have been classified into cognitively-defined subgroups based on relative impairments at the time of AD diagnosis [9], consisting of 196 as AD-Memory, 16 as AD-Executive functioning, 52 as AD-Language, 91 as AD-Visuospatial functioning, and 317 other domains (**Table S2**). Details about these cognitively-defined subgroups are presented in **Supplementary Information** and reported elsewhere [9]. We evaluated whether mbPRSs of CIH modules are linked into one of the four cognitively-defined subgroups (AD-Memory, AD-Executive, AD-Language, and AD-Visuospatial domains). Each subject was assigned into a membership of one subgroup (coded as 1), and otherwise for the other subgroups (coded as 0) without overlapping subjects between subgroups. We tested association between mbPRSs and a dichotomized membership of cognitively-defined subgroups in a logistic regression model adjusting for age, sex, the first four PCs, and genotype platform as covariates.

### 2.6. Brain imaging (MRI) data analysis with mbPRSs of the CIH modules in ADNI

To understand the relationships between our CIH modules and brain atrophy at specific locations, we tested the association between mbPRSs and surface-based cortical thickness of AD patients using general linear models after adjusting age, sex, magnetic field strength, and intracranial volume as covariates [23]. Detail information about brain imaging data processing for surface-based measure of cortical thickness in ADNI are described elsewhere [23].

### 2.7. Biological functions of genes in the CIH modules

Gene-ontology (GO) analyses were conducted to discern biological pathways of AD-associated genes in CIH modules using the Ingenuity Pathway Analysis software (QIAGEN, Redwood, CA). We also looked up associations between the CIH module-genes and AD-related neuropathological traits including Consortium to Establish a Registry for Alzheimer’s Disease (CERAD) score and Braak stage, and quantitative measures of proteins including Aβ_42_, phosphorylated Tau at 181 (pTau181) and 231 (pTau231), postsynaptic density protein 95 (PSD95), C4a, C4b, and PPP2CA/B from prefrontal cortex area of autopsied brains (FHS/BUADRC) [13].

## 3. RESULTS

### 3.1. AD-associated modules in AD brains were preserved in independent studies

Eighty-three modules were identified in the discovery dataset (FHS/BUADRC), and 29 of these modules were preserved in the two validation datasets (**Figure 1A**). Fourteen of the 29 preserved modules (M1-M14) contained genes that were significantly enriched in at least one of the AD gene-sets (NP, NFT, or AD-risk) with FDR<0.05 (**Figure 1B** and **Table 1**). Interestingly, only four modules (M1-3 and M11) were nominally enriched in the AD-risk gene-set, and all 14 modules were at least three orders of magnitude more significantly enriched in either NP or NFT gene-sets (**Table 1**). These findings may imply that our modules derived from the transcriptome datasets of AD brains (without AD-free controls) would capture the gene-sets for underlying changes in AD pathology, rather than the overall disease risk. Therefore, we linked one phenotype, either NP or NFT, but not AD-risk to each of those fourteen modules according to its most significantly enriched gene-set, and the GWAS summary data of the linked phenotype (NP or NFT) is utilized for computing each of fourteen different mbPRSs (NP-linked modules: M2, M5, M6, M10, and M13; NFT-linked modules: M1, M3, M4, M7-9, M11, M12, and M14).

**Figure 1.**
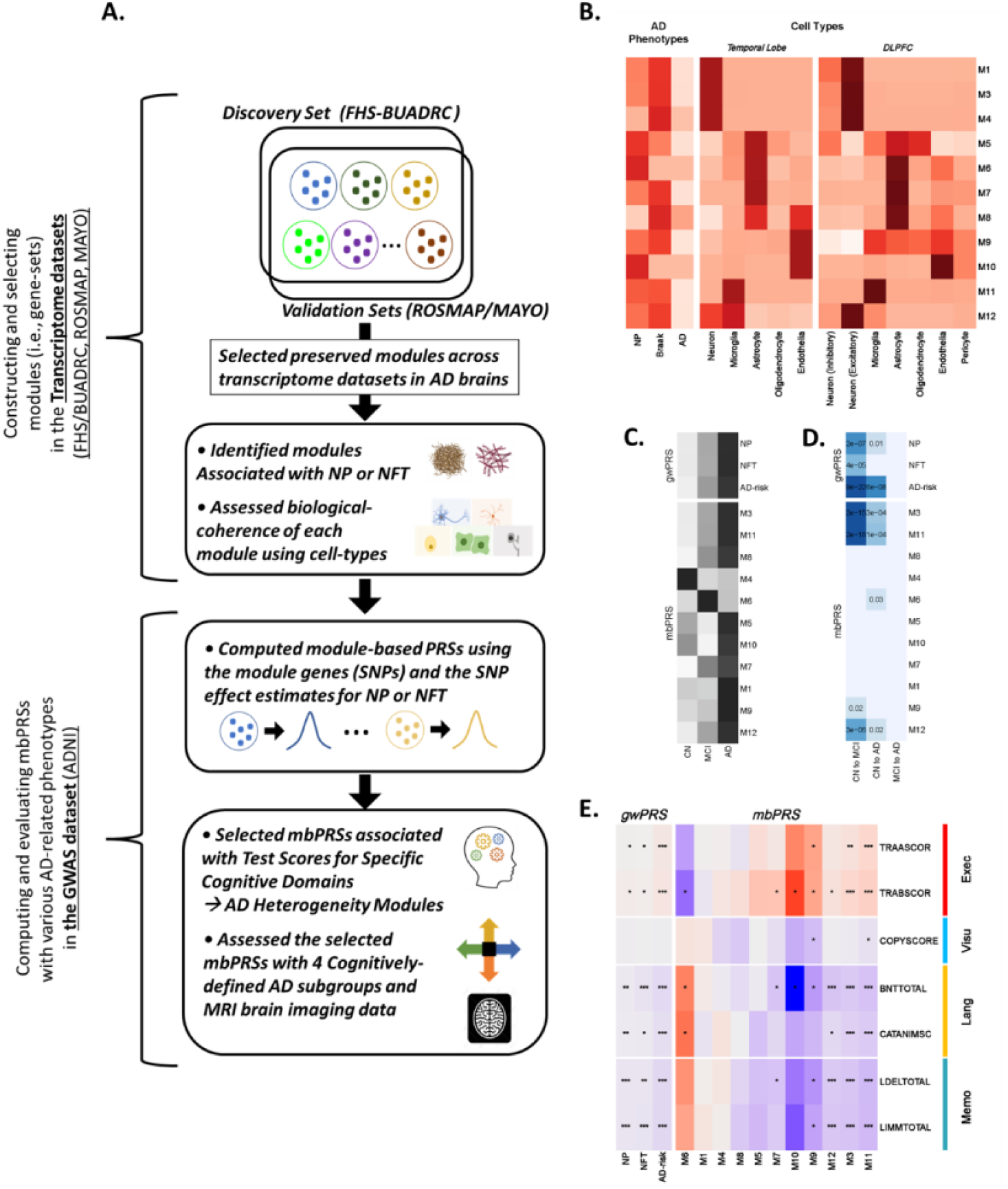
**A**. Schematic of our study design of constructing co-expression modules (sets of genes), selecting AD-associated modules, and building module-based polygenic risk scores (mbPRSs) for explaining the AD heterogeneity, which were tested and evaluated with gene-sets for AD-related neuropathological traits (NP and NFT human brain cell-types, cognitive test scores, cognitively-defined AD subgroups, and brain MRI imaging data. **B**. Enrichment strength of the eleven AD-associated modules with AD phenotypes (NP, NFT, and AD-risk) and cell type specific gene-sets in temporal lobe and dorsal lateral prefrontal cortex (DLPFC). The darker color indicates the more significant enrichment P-value. Heatmaps of **C**. mean values of PRSs across the disease stages including clinical normal (CN), mild cognitive impairment (MCI), and AD (the darker color indicates the larger mean value of the PRS.); **D**. associations between PRSs and disease progression (CN to MCI, CN to AD, MCI to AD) (the darker color indicates the more significant P-value); **E**. associations between PRSs and seven test scores for four cognitive domains (executive functioning, visuospatial functioning, language, memory). Red and blue mean positive and negative effect directions, respectively. Dots in the cells indicate the strength of associations).

**Table 1.** AD associated preserved networks in AD brains

| Module | Z-summary |  | Enrichment P-value |  |  |
| --- | --- | --- | --- | --- | --- |
|  | ROSMAP | MAYO | NP | NFT | AD-RISK |
| M1 | 40.56 | 20.71 | $3.74 \times 10^{-6}$ | $3.26 \times 10^{-9}$ | 0.04 |
| M2 | 37.45 | 20.68 | $3.50 \times 10^{-8}$ | $2.57 \times 10^{-7}$ | 0.002 |
| M3 | 35.48 | 18.59 | $5.38 \times 10^{-5}$ | $8.36 \times 10^{-7}$ | 0.01 |
| M4 | 32.32 | 18 | $3.42 \times 10^{-2}$ | $1.25 \times 10^{-5}$ | 0.07 |
| M5 | 29.83 | 15.91 | $1.46 \times 10^{-6}$ | $2.92 \times 10^{-4}$ | 0.40 |
| M6 | 26.56 | 14.46 | $4.70 \times 10^{-6}$ | 0.09 | 0.09 |
| M7 | 25.84 | 14.23 | $2.65 \times 10^{-3}$ | $2.47 \times 10^{-4}$ | 0.12 |
| M8 | 20.52 | 14.07 | 0.47 | 0.02 | 0.20 |
| M9 | 20.51 | 12.71 | $4.34 \times 10^{-3}$ | $8.89 \times 10^{-4}$ | 0.37 |
| M10 | 18.25 | 12.64 | 0.02 | 0.16 | 0.16 |
| M11 | 17.32 | 11.81 | $8.70 \times 10^{-4}$ | $7.48 \times 10^{-4}$ | 0.01 |
| M12 | 17.06 | 10.55 | 1.00 | 0.05 | 0.95 |
| M13 | 15.57 | 8.59 | 0.02 | 0.03 | 0.81 |
| M14 | 14.79 | 7.81 | 0.06 | 0.04 | 0.48 |

Module is a co-expressed gene network in the discovery from the Framingham Heart Study and Boston University Alzheimer's Disease Center (FHS/BUADRC) study.
Z-summary is a network preservation score of a module from the discovery to at least one of two validation datasets, the Religious Orders Study and Rush Memory and Aging Project (ROSMAP) and the Mayo Clinic Study of Aging (MAYO).
Enrichment p-value for a module were computed using genes in the given module containing a SNP with $P < 10^{-3}$ from *Beecham et al.* for neuritic plaque [NP] and neurofibrillary tangles [NFT] [18] and from *Kunkle et al.* for AD-risk [19].
Fourteen modules were selected when a module in the FHS/BUADRC study was preserved with $Z\text{-summary} > 5$ in both validation datasets, ROSMAP and MAYO, and significant at $p\text{-value} < 0.05$ with at least one of the gene-sets for NP, NFT, or AD-risk.

All 14 AD-associated modules were significantly enriched in specific cell-types, where these results were consistent between temporal lobe and prefrontal cortex regions (**Figure 1B** and **Table S3**). M1 to M4 were predominantly enriched in excitatory neurons (best P with M2 from prefrontal cortex=4.4×10^−188^), M6 and M7 in astrocytes (best P with M7 from prefrontal cortex =1.3×10^−97^), M10 in endothelia (P from temporal lobe=9.9×10^87^), and M11 in microglia (P from temporal cortex=1.7×10^−129^). The other five modules (M5, M8, M9, M12 and M13) were significantly enriched in more than one cell type, while M5 and M8 (astrocytes), M9 (endothelia), M12 (neurons), and M13 (microglia) were significantly enriched in at least one cell type in both brain regions.

### 3.2. Module-based PRSs explained heterogeneity in cognitive functions among AD Patients

For each of 14 modules, we extracted module-SNPs and their effect estimates on the module-assigned phenotype for computing mbPRS in the ADNI GWAS sample. Of the 14 modules, we computed mbPRSs of the nine NFT-linked modules (M1, M3, M4, M7-9, M11, M12, and M14) using effects of module SNPs on NFT, while mbPRSs of the remaining five NP-linked modules (M2, M5, M6, M10, and M13) using NP. Three modules including M2, M13, and M14 were excluded due to their low standard error (<0.05) and/or their extremely skewed distributions for the following analyses (**Table S4**).

In comparison, we observed that three gwPRSs were significantly correlated with mbPRSs of the M3, M11, and M12 modules (correlation r^2^≥0.1), while the rest eight mbPRSs were not correlated with each other (r^2^<0.01). The mean values of all three gwPRSs were sequentially increased from CN to MCI and AD (**Figure 1C**). In contrast, the mean values of mbPRS were varied across the disease stages, which the mean values of modules (M3-6, and M10) were smaller in MCI or AD stages than in the CN stage (**Figure 1C**). For the disease progression, two gwPRSs (NFT and AD-risk) and three mbPRSs (M3, M11, and M12) were significantly associated with the progressions from CN to both MCI and AD (**Figure 1D**). None of PRSs were associated with the progression from MCI to AD. Interestingly, NFT-gwPRS and M9-mbPRS were associated with the progression from CN to MCI, and M6-mbPRS was associated with the progression from CN to AD. All three gwPRSs were significantly associated with most cognitive test scores, except for the visuospatial domain (COPYSCORE), with the consistent effect directions across cognitive tests. This indicates that the gwPRSs are not likely to differentiate cognitive impairment heterogeneity among AD cases (**Figure 1E**).

Five mbPRSs from M3, M6, M9, M11, and M12 were robustly associated (P-value<0.05) with at least two cognitive domains, while five mbPRSs from M1, M4, M5, M8, and M10 showed no association (P-value>0.05) with any cognitive test scores (**Figure 1E** and **Table S5**). Of the 5 mbPRSs with robust associations for cognitive domains, mbPRSs of M3 and M11 were strongly associated with the 3 cognitive domains except for the visuospatial functioning, indicating that mbPRSs from M3 and M11 would not differentiate the cognitive impairment heterogeneity. The M6-mbPRS was nominally associated with all two language-domain test scores (BNTTOTAL P-value=0.03 and CATANIMSC P-value=0.01). Although the M9-mbPRS was associated with cognitive test scores of multiple domains, the M9-mbPRS showed association with the visuospatial functioning (COPYSCORE P-value=0.05), which is distinctive across all pairs of associations between PRS and cognitive scores. The M12-mbPRSs were strongly associated with language (best P with BNTTOTAL=2.2×10^−6^) and memory (best P with LIMMTOTAL=4.4×10^−6^) domains (**Table 2**). Therefore, we prioritized M6, M9, and M12 as CIH modules and attempted to validate the associations between mbPRSs of the CIH modules (M6, M9, and M12) and the cognitive test scores among the AD cases in FHS (**Table S6**). We observed nominally significant associations (P<0.05) between the M6-mbPRS and two language-domain cognitive test scores in the FHS AD cases (BNT30 P-value=0.03 and BNT30cue P-value=0.03; **Table S7**). Although we did not find associations of the rest two modules (M9 and M12) with the cognitive test scores in FHS, the three CIH modules (M6, M9, and M12) were further tested with cognitively-defined subgroups [9] and brain atrophy among AD patients.

**Table 2.** Associations between cognitive test scores and three module-based PRSs

| Cognitive Domain | Cognitive Test | M6 |  |  | M9 |  |  | M12 |  |  |
| --- | --- | --- | --- | --- | --- | --- | --- | --- | --- | --- |
|  |  | BETA | SE | P-value | BETA | SE | P-value | BETA | SE | P-value |
| Executive functioning | TRAASCOR | -0.16 | 0.12 | 0.17 | 0.18 | 0.06 | $1.08 \times 10^{-3}$ | 0.02 | 0.02 | 0.15 |
| | TRABSCOR | -0.26 | 0.12 | 0.03 | 0.18 | 0.06 | $1.58 \times 10^{-3}$ | 0.03 | 0.02 | 0.05 |
| Visuospatial functioning | COPYSCORE | 0.03 | 0.12 | 0.79 | -0.12 | 0.06 | 0.049 | -0.01 | 0.02 | 0.77 |
| Language | BNTTOTAL | 0.26 | 0.12 | 0.03 | -0.17 | 0.06 | $2.57 \times 10^{-3}$ | -0.08 | 0.02 | $2.23 \times 10^{-6}$ |
|  | CATAANIMSC | 0.29 | 0.12 | 0.01 | -0.11 | 0.06 | 0.05 | -0.04 | 0.02 | 0.01 |
| Memory | LDELTOTAL | 0.22 | 0.12 | 0.06 | -0.18 | 0.06 | $1.14 \times 10^{-3}$ | -0.07 | 0.02 | $4.55 \times 10^{-5}$ |
| | LIMMTOTAL | 0.18 | 0.12 | 0.12 | -0.15 | 0.06 | $7.18 \times 10^{-3}$ | -0.08 | 0.02 | $4.42 \times 10^{-6}$ |

LIMMTOTAL and LDELTOTAL: logical memory immediate and delayed recall tests; TRAASCOR and TRABSCOR: trail marking test A/B. CATANIMSC: category fluency animal score. BNTTOTAL: Boston naming test total. COPYSCORE: clock test total score
Beta estimates (BETA), standard error (SE), and P-values were calculated with module-based PRSs of the M2, M6, and M9 modules for domain specific cognitive functions.

### 3.3. Module-based PRS associations with cognitively-defined AD subgroups and brain atrophy

Of the three CIH mbPRSs, mbPRSs of M6 and M9 showed nominal associations at P<0.05 with odds ratio (OR)>1.0 with the cognitively-defined subgroups for AD-Language (OR=5.5; P=0.01) and AD-Visuospatial functioning (OR=1.9; P=0.04), respectively (**Table 3**), but M12-mbPRS was associated with none of subgroups. In contrast, the gwPRSs for NP and AD-risk failed to differentiate any of the subgroups, and only NFT-gwPRS was nominally associated with the AD-Memory subgroup (OR=1.01; P=0.04).

**Table 3.** Membership assignment of three module-based PRSs in four cognitively-defined AD subgroups in ADNI

| Cognitively-defined Subgroups | gwPRS |  |  |  |  |  | mbPRS |  |  |  |  |  |
| --- | --- | --- | --- | --- | --- | --- | --- | --- | --- | --- | --- | --- |
|  | NP |  | NFT |  | AD-Risk |  | M6 |  | M9 |  | M12 |  |
|  | OR (95% CI) | P | OR (95% CI) | P | OR (95% CI) | P | OR (95% CI) | P | OR (95% CI) | P | OR (95% CI) | P |
| **Executive Functioning** | 0.99 (0.93-1.0) | 0.69 | 1.00 (0.91-1.10) | 0.98 | 1.01 (0.93-1.1) | 0.65 | 0.37 (0.04-3.60) | 0.39 | 0.95 (0.34-2.60) | 0.92 | 1.10 (0.78-1.5) | 0.66 |
| **Language** | 0.99 (0.95-1.0) | 0.43 | 0.99 (0.93-1.01) | 0.74 | 0.97 (0.91-1.0) | 0.24 | **5.50 (1.5-20.0)** | **0.009** | 0.81 (0.42-1.50) | 0.51 | 0.92 (0.75-1.1) | 0.40 |
| **Memory** | 1.00 (0.99-1.0) | 0.57 | **1.01 (1.00-1.11)** | **0.04** | 1.00 (1.0-1.1) | 0.08 | 0.97 (0.44-2.2) | 0.95 | 1.20 (0.84-1.80) | 0.29 | 1.00 (0.92-1.2) | 0.56 |
| **Visuospatial Functioning** | 1.00 (0.97-1.0) | 0.75 | 0.96 (0.91-1.01) | 0.09 | 0.96 (0.91-1.0) | 0.07 | 0.88 (0.28-2.8) | 0.83 | **1.91 (1.11-3.31)** | **0.04** | 1.00 (0.87-1.2) | 0.76 |

OR: odds ratio. CI: confidence interval. Cognitively-defined AD subgroups in ADNI have been previously defined [9]. Bold values with significance at $P < 0.01$ and $OR > 1.0$ indicate these modules are likely members of the corresponding cognitively-defined subgroups.

We observed that mbPRSs of M6 and M9 were significantly associated with cortical thickness at specific brain locations (M6: bilateral frontal, parietal, and temporal lobes; M9: bilateral frontal lobes; **Figure 2A**). Interestingly, higher M6-mbPRS and lower M9-mbPRS, which were associated with better cognitive functions in our study, were significantly associated with larger cortical thickness, which represent less brain atrophy (**Figure 2A**). Particularly, the M6-mbPRS was strongly linked to the brain atrophy at the Wernicke area where lesions have been associated with severe impairments of word comprehension [24].

**Figure 2.**
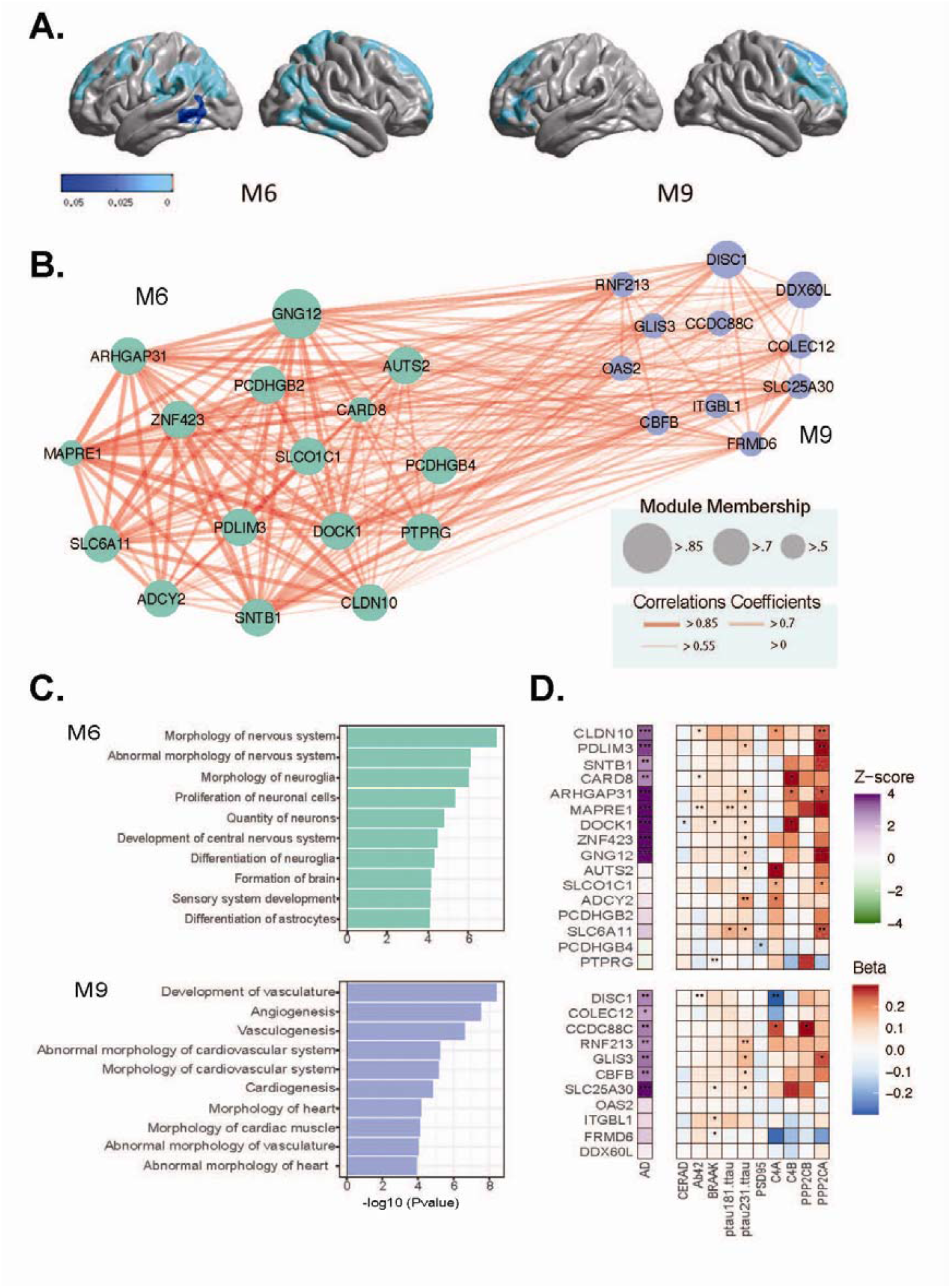
M6 and M9 were selected as cognitive impairment heterogeneity (CIH) modules. **A**. P-value map (threshold at p<0.05; the darker blue color indicates significant P-value.) showing association between cerebral cortical thickness and module-based polygenic risk scores for M6 and M9. **B**. Co-expression network of genes in M6 and M9. **C**. Biological functions (gene ontology) of M6 and M9. **D**. Associations of the expression levels of genes in M6 and M9 with AD risk and AD-related biomarkers (previously published [13]).

### 3.4. Functional profiling of M6 and M9

Among the module-genes in M6 and M9, we focused on the GWAS genes (*i*.*e*., seed genes) with a SNP with P<0.001 for NP or NFT (# of GWAS genes, M6=16 and M9=11; **Figure 2B** and **Table S8**). The seed genes in M6 were significantly enriched in pathways (**Figure 2C** and **Table S9**) including morphology of nervous system (P=4.0×10^−8^), abnormal morphology of nervous system (P=7.8×10^−7^) and differentiation of astrocytes (P=8.4×10^−5^), and the M9 seed genes were in vascular system including development of vasculature (P=3.8×10^−9^), angiogenesis (P=3.0×10^−8^), and vasculogenesis (P=2.5×10^−7^) (**Figure 2C** and **Table S8**).

According to our previous report [13], the majority of the seed genes in M6 (best: *DOCK1*, P=3.0×10^−7^) and M9 (best: *SLC25A30*, P=6.3×10^−6^) were up-regulated in AD compared with control brains (**Table S10**). In addition, we observed significant associations between expression levels of the seed genes in M6 and M9 and AD-related protein levels (**Figure 2D** and **Table S10**). The seed genes in M6 were significantly associated (P<0.05) with CERAD scores, Braak stages, Aβ_42_, pTau_181_/tTau ratio, pTau_231_/tTau ratio, C4a, C4b, and PSD95 (**Table S11**), which the most significant association was observed with expression of *ADCY2* with pTau_181_/tTau ratio (P-value=1.1×10^−3^). The expressions of the seed genes in M9 were nominally significant with Braak stages, Aβ_42_, pTau_231_/tTau ratio, and C4a levels with the best P-value between expression of *DISC1* and C4a (P-value=1.0×10^−3^).

## 4. DISCUSSION

### 4.1. Key findings

The goals of this study were to identify gene-sets responsible for the biological mechanisms underlying AD heterogeneity and to test use of genetic information (*e*.*g*., PRS) derived from the gene-sets for prediction of individual risks to clinical AD-subgroups. We generated modules (gene-sets) that were commonly observed in multiple transcriptome datasets of AD brains. We closely evaluated biological coherence (*e*.*g*., cellular identifies) and disease relevance of the modules using profiling of human brain cell types and genetics of AD neuropathology. Then, we selected the CIH modules (M6, M9, and M12), which are likely to explain the disease heterogeneity in cognitive impairment of the AD patients, by testing with domain-specific cognitive test scores in ADNI (clinic-based study), and association with M6-mbPRS was replicated in FHS (population-based study). This demonstrates the concept we proposed in the current manuscript can be generalizable and applicable to diverse populations, although not all the modules are available in all populations. We attempted to validate whether those CIH modules differentiate cognitively-defined AD subgroups and found that the two CIH modules (M6 and M9), which showed associations with cognitive test scores for language and visuospatial domains, respectively, markedly recognized the matching AD subgroups (AD-Language and AD-Visuospatial). Furthermore, we found that those CIH modules (M6 and M9) were also linked to atrophy in specific brain areas (M6: Wernicke’s area in temporoparietal cortex; M9: frontal cortex) which were previously reported to underpin for language comprehension [24, 25] and visuospatial deficit [26].

### 4.2. Advantage of mbPRSs for AD subgrouping

The three gwPRSs for NP, NFT, and AD-risk showed high correlations each other and largely similar patterns from associations with disease conversion and cognitive test scores. In contrast, our mbPRSs showed almost no correlations each other and are associated with the disease progression at certain stages and performance of specific cognitive domains. These findings after comparing our novel mbPRSs and traditional gwPRSs demonstrate that mbPRSs would be more useful for explaining the phenotypic heterogeneity in AD patients, while gwPRSs (*i*.*e*., traditional PRS) would be more relevant to predict the overall disease risk. Our mbPRSs successfully distinguish differences in clinical (cognitive domains) and structural brain imaging patterns, indicating representation of different disease mechanisms and thereby would be effective tools for dissecting the disease heterogeneity. The gwPRSs for NP and AD-risk failed to recognize the AD subgroups. Only the gwPRS for NFT discerned the most typical cognitive subgroup, AD-Memory domain. However, newly identified mbPRSs for the M6 an M9modules recognized different types of AD subgroups. This indicates that the conventional gwPRS approach is less likely to recognize differences between AD subtypes. Further, these results support our hypothesis that subgrouping genetic markers from gene-sets responsible for a distinct disease mechanism leading to an AD subtype is important for precision medicine and genome-guided clinical trials.

There have been huge efforts to improve predicting and distinguishing disease subtypes using polygenic profiling for early detection of subjects at risk [27-29]. Polygenic risk scores can be also useful to predict expected development of a disease or treatment responses in particular patient subgroups [30]. Our module-based polygenic profiling has innovative features compared to those previously conducted co-expression studies [11, 12] and conventional PRS approaches for AD [29-32]. First, our co-expression modules were developed from only AD brains excluding CN and MCI brains, while previous co-expression studies used transcriptome data of AD cases together with controls [11, 12]. Biological processes underlying disease heterogeneity in AD brains may be different from CN or MCI brains [33, 34]. Inclusion of non-AD transcriptome data would well differentiate gene-sets relevant to the disease risk but not explain disease heterogeneity. Second, previous polygenic profiling studies have generated PRSs by aggregating genetic estimates of genome-wide or most significant SNPs, which may have improved prediction rates [30] but cannot explain specific biological functions. In contrast, our mbPRSs are derived from biologically coherent gene-sets, which enable us to interpret biological functions of the modules and thereby provide insights on functional/mechanistic pathways for the AD subtypes. A previous study demonstrated genomic annotations at the single tissue level can improve our understanding on the etiology of complex human diseases [35]. A recent simulation study with failed AD trials confirms that the main failure reason is because variability between individuals’ in trials masks efficacy [3]. Therefore, our mbPRSs relevant to cell/tissue-level transcriptome profiles, brain imaging data, and cognitively-defined subgroups can be utilized for studying disease subtypes, prognosis, and response of treatment.

### 4.3. Role of omics and genetic profiling in AD subgrouping

Profiling using the omics data including transcriptome data at the tissue- or the cell-level helped identify clinically and neuropathologically heterogeneous modules but also understand the biological functions of the modules. For example, the identified M6 module-genes were enriched for astrocytes, neuritic plaque scores, and language domain of cognitive function. This confirms the previous report that astrocytes are involved in amyloid clearance [36] and damaged astrocytes impact language domain among AD patients [37]. Our discovery showed that M9 module-genes are linked to endothelial cells, Braak stages, and visuospatial functioning in this study. Increased vascular inflammation in endothelial cells has been observed among AD patients with poor short-term visuospatial functioning [38].

Genes in the M6 and M9 modules have been previously reported for association with neurodegenerative diseases. Most of genes in the two modules have biological functions relevant to the nervous system or have been previously reported in genetic or experimental studies for neurodegenerative diseases. For example, *SLC6A11* in M6 has been targeted for drug development of different neurodegenerative diseases including epilepsy [39]. *GLIS3* in M9 has been associated with T2D [40, 41] and a longer life expectancy [42]. SNPs from *GLIS3* in M9 showed genome-wide significant associations from GWASs for amyloid-β and phosphorylated tau proteins in cerebrospinal fluid (CSF) [43].

### 4.4. Limitations

Our study has several limitations. First, the sample size of discovery AD brains was modest. Therefore, we did not have statistical power for explaining the subtle phenotypic variations among AD patients, which might lead to detect modules associated with a few specific cognitive domains. In addition, our current study exclusively relied on cognitive test scores for prioritizing CIH modules, which may not be useful for detecting unknown or brain imaging-based subtypes of the disease. We also acknowledge that our findings in ADNI may not represent AD heterogeneity in other populations. However, since one of modules was replicated in an independent study (FHS), there are shared mechanisms across diverse populations. Since we focused on AD patients, our sample size of AD subgroups remained underpower, so we could not apply multiple testing correction in current study. This limitation was mitigated by replicating one of the mbPRSs in FHS. Second, we could apply and attempt to validate our module-based approach in other AD GWAS sample with different types of AD subgroups based on CSF biomarker [44] or brain imaging data [1], but the sample sizes of those datasets available to the public will be extremely limited. Third, most of previously defined cognitive subgroups in ADNI were predominantly classified as subgroups of memory (31.8%), while subgroups for executive functioning (2%) was relatively limited, which may lead to no significant associations between PRSs and uncommon subgroups (e.g., AD-Executive). Moreover, it will be necessary to repeat analyses in independent samples to validate our findings as well as our approach. In general, the datasets with GWAS for enough AD patients with carefully classified clinical phenotypes and clinically and/or pathologically defined subtypes were extremely limited. However, efforts to subgroup AD cases in additional datasets are ongoing. Also, the GWAS summary statistics for AD neuropathological traits (NP and NFT) in this study were generated based on genotype imputation using a previous reference panel (1000 genome) [18], which may affect quality and accuracy of our gene-sets and PRSs. However, we used common SNPs (MAF>5%) for constructing gene-sets and PRSs, and the imputation qualities of common SNPs are still relatively acceptable even in the previous reference panel [45]. Therefore, potential problems caused by low imputation quality would be largely limited in our study.

Future work in other independent GWAS sample with cognitively-defined subgroups (or relevant subgroups based on cognitive tests) will be required to validate our module-based prediction of AD subtypes. Furthermore, linking genetics of various disease-related phenotypes to the target disease would enhance to dissect the disease heterogeneity [10]. Other AD-related GWAS summary data including cerebral amyloid angiopathy, hypertension, cholesterol, and insulin resistance can be added for extending AD phenotype gene-sets, which will lead us to detect novel gene-sets and to recognize other subgroups beyond AD-Language/Visuospatial domains.

### 4.5. Conclusion

In conclusion, PRSs developed using biologically coherent gene-sets and disease-related phenotypes can successfully differentiate cognitively-defined subgroups and brain region specific atrophy, which likely represent mechanistic pathways responsible for disease subtypes. Classification of patients using genetic information will allow patient subgrouping and target prioritization for the subgroups, which may eventually lead to precision medicine in AD. Our study warrants in extensive validations in large datasets.

## Data Availability

FHS data are available on the dbGaP (Study Accession ID: phs000056.v5.p3). ROSMAP resources can be requested at from the CommonMind Consortium portal (http://www.synapse.org). Data used in preparation of this article were obtained from the Alzheimer Disease Neuroimaging Initiative (ADNI) database (http://adni.loni.usc.edu). As such, the investigators within the ADNI contributed to the design and implementation of ADNI and/or provided data but did not participate in analysis or writing of this report. A complete listing of ADNI investigators can be found at: http://adni.loni.usc.edu/wp-content/uploads/how_to_apply/ADNI_Acknowledgement_List.pdf.

https://www.niagads.org/

## Data Availability Statement

Genotypes and clinical/neuropathological phenotype data are accessible by directly applying to the LONI portal for the ADNI at http://adni.loni.usc.edu. Summary statistics for GWAS studies can be accessed by applying directly to the National Institute on Aging Genetics of Alzheimer’s Disease Data Storage Site (NIAGADS), a NIA/NIH-sanctioned qualified-access data repository, under accession <u>NG00075</u>. Data supporting the findings of this study are available from the NIAGADS website (https://www.niagads.org/).

## Acknowledgements

This study was supported by the National Institute on Aging (NIA) grants, U01-AG068057, U19-AG068753, P30-AG072978, and RF1-AG057519. GWAS summary statistics used in this study were distributed by the National Institute on Aging Alzheimer’s Disease Data Storage Site (NIAGADS) at the University of Pennsylvania (U24-AG041689-01). Collection of study data provided by the Rush Alzheimer’s Disease Center, Rush University Medical Center, Chicago was supported through funding by NIA grants P30-AG10161, R01-AG15819, R01-AG17917, R01-AG30146, R01-AG36836, U01-AG32984, U01-AG46152, and U01-AG61358, and funding from the Illinois Department of Public Health and the Translational Genomics Research Institute. Study data were also provided by Dr. Nilüfer Ertekin-Taner and Dr. Steven G. Younkin, Mayo Clinic, Jacksonville, FL using samples from the Mayo Clinic Study of Aging, the Mayo Clinic Alzheimer’s Disease Research Center, and the Mayo Clinic Brain Bank. Collection of these data was supported through funding by NIH grants P50-AG016574, R01-AG032990, U01-AG046139, R01-AG018023, U01-AG006576, U01-AG006786, R01-AG025711, R01-AG017216, R01-AG003949, and R01-NS080820, and by funding from the CurePSP Foundation and the Mayo Foundation. Data collection and sharing for this project was funded by the Alzheimer’s Disease Neuroimaging Initiative (ADNI) (National Institutes of Health Grant U01 AG024904) and DOD ADNI (Department of Defense award number W81XWH-12-2-0012). ADNI is funded by the National Institute on Aging, the National Institute of Biomedical Imaging and Bioengineering, and through generous contributions from the following: AbbVie, Alzheimer’s Association; Alzheimer’s Drug Discovery Foundation; Araclon Biotech; BioClinica, Inc.; Biogen; Bristol-Myers Squibb Company; CereSpir, Inc.; Cogstate; Eisai Inc.; Elan Pharmaceuticals, Inc.; Eli Lilly and Company; EuroImmun; F. Hoffmann-La Roche Ltd and its affiliated company Genentech, Inc.; Fujirebio; GE Healthcare; IXICO Ltd.; Janssen Alzheimer Immunotherapy Research & Development, LLC.; Johnson & Johnson Pharmaceutical Research & Development LLC.; Lumosity; Lundbeck; Merck & Co., Inc.; Meso Scale Diagnostics, LLC.; NeuroRx Research; Neurotrack Technologies; Novartis Pharmaceuticals Corporation; Pfizer Inc.; Piramal Imaging; Servier; Takeda Pharmaceutical Company; and Transition Therapeutics. The Canadian Institutes of Health Research is providing funds to support ADNI clinical sites in Canada. Private sector contributions are facilitated by the Foundation for the National Institutes of Health (www.fnih.org). The grantee organization is the Northern California Institute for Research and Education, and the study is coordinated by the Alzheimer’s Therapeutic Research Institute at the University of Southern California. ADNI data are disseminated by the Laboratory for Neuro Imaging at the University of Southern California.

