## Supplementary Materials for "Alzheimer’s Disease Heterogeneity Explained by Polygenic Risk Scores Derived from Brain Transcriptomic Profiles"

**Supplementary Information**

**RNA-Seq data of AD brains from Framingham Heart Study (FHS) and Boston University Alzheimer’s Disease Research Center (BUADRC)**

The sequence reads with low quality scores were removed with FASTX-Toolkit and trimmomatic [1]. The processed reads were mapped onto reference genome GRCh38 with Bowtie [2]. The expression values were estimated in TPM (Transcripts per million) by RSEM [3]. Genes with low expression after log-transformation (mean < 2.0) were filtered out for further analysis [4]. After the rigorous QC [4], the expression data with 24,358 unique genes of 65 AD patients was used for constructing co-expression network model in dorsal lateral frontal cortex (DLPFC) region of 65 AD brains in FHS/BUADRC.

The AD diagnosis of subjects in BUBB/FHS was proceeded based on criteria for possible, probable, or definite AD from the National Institute of Neurological and Communicative Disorders and Stroke and the Alzheimer’s Disease and Related Disorders Association (NINCDS–ADRDA) [5, 6].

**Constructing the coexpression Network Model in the RNS-Seq gene expression data of BUBB/FHS, ROSMAP, and MAYO Clinic**

We constructed coexpression network models using RNA-Seq gene expression data of AD cases from three independent datasets including FHS/BUADRC [4], ROSMAP [7], and MAYO Clinic [8]. We downloaded the version #1 data files for RNA-Seq data of both ROSMAP and MAYO Clinic, which were deposited in 2015 at the Synapse.org. We used the software, weighted gene coexpression network analysis (WGCNA) for constructing the network models [9]. This software define module of highly coexpressed (i.e., coregulated) genes, which are computed by Pearson correlation analysis of gene expression levels across sample. For constructing the models, we applied 12 for the parameter value β (power), which is for reducing the effects of weak correlation and increasing effect of strong correlation among the genes. Co-expression modules were defined from the hierarchical tree by “dynamic tree cut algorithm” [9]. The modules including less than 20 genes were considered as noisy modules and ignored for the further analyses.

Preservation of the discovery modules was evaluated in the two independent validation datasets, including ROSMAP and MAYO Clinic, using z-summary statistics [9]. The z-summary statistics are the composite scores derived from comparison of density and connectivity of modules from the discovery and validation datasets. We considered a module to be preserved if z-summary scores greater than 5.0 in both validation datasets [9].

**Defining the gene-sets for human brain cell types**

Single cell RNA-Seq data from the temporal lobe brain region: We used single cell RNA-Seq data of human brain cells published by *Darmanis et al*. [10]. the mapped read counts, which were normalized into CPM (count per million), were downloaded from Gene Expression Omnibus (Accession number: GSE67835). Cell type-specific genes were defined when median and mean expression levels (CPM) of genes greater than 4.0.

Single nucleus RNA-Seq data from dorsal lateral frontal cortex: We previously generated cell type gene-sets using the single nucleus RNA-Seq data of human brain dorsal lateral frontal cortex in the ROSMAP sample [4, 11]. Briefly, raw FASTQ single-nucleus RNA sequencing data of 48 subjects (24 AD cases and 24 controls) in the ROSMAP dataset were obtained from the Synapse database. Read counts were aligned to a reference genome (GRCh38) by CellRanger software (v.3.1.0). We conducted principal component (PC) analysis to reduce the model complexity of the high variable expression of 3,179 genes (marker genes). A cell type-specific expression metric was calculated by dividing the expression in each cell type by the total expression across all cell types. The top 10 PCs were included in t-SNE analysis. For each cluster, cell-type labels were assigned by known marker genes [11], which yielded seven cell type-specific clusters representing astrocytes, endothelial cells, excitatory neurons, inhibitory neurons, microglia, oligodendrocytes, and pericytes.

**Subjects and GWAS Data in in the Alzheimer’s Disease Neuroimaging Initiative (ADNI)**

The ADNI is a longitudinal study assessing clinical, imaging, genetic, and biospecimen biomarkers from US-based subjects in various disease stages including cognitively normal (CN), mild cognitive impairment (MCI), and AD. We downloaded genetic data and AD-related domain-specific cognitive test scores of the ADNI sample from the LONI website (<http://adni.loni.usc.edu>). The genetic data of ADNI samples from two different arrays (ADNI-1, n=679 and ADNI-GO/2, n=397) were cleaned and imputed using the Haplotype Reference Consortium reference panel were performed.

**Quality Control of GWAS Datasets in ADNI**

The ADNI-1 and ADNI-GO/2 samples were genotyped using the Illumina Human610-Quad, and HumanOmniExpress microarray chips (Illumina, Inc., San Diego, CA), respectively. QC procedures for the genotype data were performed using PLINK [12]. Samples with genotyping call rates<95% were excluded. Subjects with non-European ancestry were removed based on the self-reported ethnicity. SNPs were filtered out where genotype missingness were greater than 5%, Hardy-Weinberg equilibrium P-value was less than 10^-6^, and minor allele frequency (MAF) is less than 5%, and subjects with missing genotype > 5% were excluded. Population substructures in subjects from two ADNI genotype chips were separately evaluated using the smartpca script in EIGENSTRAT software [13]. A total of 548,010 SNPs for ADNI-1 and 641,106 SNPs for ADNI-GO/2 were retained after cleaning. Genome-wide imputation of SNP allele dosages was conducted using Haplotype Reference Consortium reference panel.

Population substructure was evaluated in each dataset with reference populations from the 1000 Genome Project by principal components analysis using EIGENSTRAT [13]. Only individuals clustering with European samples were retained for further analysis. Population substructure in the remaining European ancestry sample was assessed using genotyped SNPs that passed QC. These SNPs were pruned to remove pairs with high linkage disequilibrium (LD) based on pairwise LD (r2) > 0.2 using PLINK and using a window size of 1,500 SNPs. Principal components (PCs) of ancestry were derived using the smartpca script in EIGENSTRAT.

**Computing Polygenic Risk Scores (PRSs) for the AD Associated Modules in ADNI**

We computed mbPRSs of the genes in the selected AD-associated modules using the following strategy. First, we choose one GWAS summary statistics (among NP, NFT[14], and AD risk[15]) for each AD-associated module according to the corresponding AD phenotype, which showed the most significant enrichment strength from the Fisher’s exact test. Second, we selected SNPs located within the module-genes with by P-value<0.001 from the chosen GWAS summary. Third, we computed mbPRS for each module using the effect estimates of the SNPs from the chosen GWAS summary. For computing genome-wide PRS (gwPRSs), we selected SNPs by P-values (<0.001) from the chosen GWAS summary.

SNPs were filtered out by multiallelic/ambiguous polymorphism, minor allele frequency (<5%), imputation quality (R^2^<0.4) and were further pruned by linkage disequilibrium (r^2^<0.4). Then, we calculated PRSs of individuals in the ADNI by sum of the imputed dosages of minor alleles weighted by the selected SNPs’ effect sizes estimated from the chosen GWAS summary statistics.

**AD Sample in Framingham Heart Study (FHS)**

The Framingham Heart Study is a multi-generation community-based family study which initiated for understanding cardiovascular disease in 1948 among 5209 individuals comprising the original cohort [16]. Participants in FHS were genotyped at Affymetrix (Santa Clara, CA) using the Affymetrix GeneChip® Human Mapping 500K Array Set. Gnome-wide association study (GWAS) data including raw genotyping calls were obtained from the dbGaP website ([http://www.ncbi.nlm.nih.gov/gap. Accessed 2012 July 2](http://www.ncbi.nlm.nih.gov/gap.%20Accessed%202012%20July%202); dbGaP Study Accession ID: phs000056.v5.p3).

We applied the same QC procedures used in the ADNI for the FHS GWAS sample. Genome-wide imputation of SNP allele dosages was conducted using HRC reference panel. We computed mbPRSs of the three CIH modules (M6, M9, and M12) for the AD cases of the FHS using the same procedure used in the ADNI sample. Subjects in FHS were administered a neuropsychological test battery using standard protocols and trained examiners, which includes trail making test A/B (TRAA and TRAB) for executive functioning, visual reproductions Immediate and Delayed Recalls (VRI and VRD) for visuospatial functioning, Boston naming test (BNT30) and with cue (BNT30cue) for language, and logical memory immediate and delayed recalls (LMI and LMD) for memory. Details of the tests administered and normative values for the FHS have been published [17, 18]. Cognitive test scores measured at the last visits were analyzed in this study. In this study, we analyzed 134 AD cases (age > 65) in FHS with GWAS and cognitive test score datasets. The sample demographic information about the FHS AD sample is as follows.

**Clinically-defined AD subgroups**

Previously, *Mukherjee et al*. [19] computed average scores (0 ~ 4) of the four cognitive domains based on individual cognitive test scores and classified the AD patients based on the scores into six potential subgroups (executive functioning, language, memory, visuospatial functioning, no and multiple domains). The first four subgroups (executive functioning, language, memory, visuospatial functioning domains) includes the AD patients with an isolated substantial relative impairment in one of four domains. The “AD-No domain” subgroup includes those with no domain with a substantial relative impairment (< 0.80 standard deviation units), and the “AD-Multiple domain” subgroup is for those with multiple domains with substantial relative impairments (> 0.80).

**Table S1**. Sample demography of AD-related cognitive tests in ADNI

| **Cognitive Test** | **Subgroup** | **N** | **Females** | **Age** |
| --- | --- | --- | --- | --- |
| LDELTOTAL | ALL | 1122 | 476 | 74.3 (7.1) |
|  | CN | 288 | 134 | 73.7 (6.4) |
|  | MCI | 370 | 149 | 74 (7.5) |
|  | AD | 464 | 193 | 74.9 (7.2) |
| LIMMTOTAL | ALL | 1122 | 476 | 74.3 (7.1) |
|  | CN | 288 | 134 | 73.7 (6.4) |
|  | MCI | 370 | 149 | 74 (7.5) |
|  | AD | 464 | 193 | 74.9 (7.2) |
| TRAASCOR | ALL | 1122 | 476 | 74.3 (7.1) |
|  | CN | 288 | 134 | 73.7 (6.4) |
|  | MCI | 370 | 149 | 74 (7.5) |
|  | AD | 464 | 193 | 74.9 (7.2) |
| TRABSCOR | ALL | 1122 | 476 | 74.3 (7.1) |
|  | CN | 288 | 134 | 73.7 (6.4) |
|  | MCI | 370 | 149 | 74 (7.5) |
|  | AD | 464 | 193 | 74.9 (7.2) |
| CATANIMSC | ALL | 1122 | 476 | 74.3 (7.1) |
|  | CN | 288 | 134 | 73.7 (6.4) |
|  | MCI | 370 | 149 | 74 (7.5) |
|  | AD | 464 | 193 | 74.9 (7.2) |
| BNTTOTAL | ALL | 1122 | 476 | 74.3 (7.1) |
|  | CN | 288 | 134 | 73.7 (6.4) |
|  | MCI | 370 | 149 | 74 (7.5) |
|  | AD | 464 | 193 | 74.9 (7.2) |
| COPYSCORE | ALL | 1122 | 476 | 74.3 (7.1) |
|  | CN | 288 | 134 | 73.7 (6.4) |
|  | MCI | 370 | 149 | 74 (7.5) |
|  | AD | 464 | 193 | 74.9 (7.2) |

LIMMTOTAL and LDELTOTAL: logical memory immediate and delayed recall tests; TRAASCOR and TRABSCOR: trail marking test A/B. CATANIMSC: category fluency animal score. BNTTOTAL: Boston naming test total. COPYSCORE: clock test total score

**Table S2.** Sample demography of cognitively defined AD subgroups in ADNI

| **Subgroup** | **N** | **Females** | **Age** |
| --- | --- | --- | --- |
| Executive functioning | 16 | 5 | 74.1 (8.0) |
| Language | 52 | 10 | 75.8 (6.1) |
| Memory | 196 | 64 | 75.3 (6.7) |
| Visuospatial functioning | 91 | 21 | 73.1 (9.4) |
| Multiple domain | 28 | 5 | 74.2 (7.4) |
| No domain | 289 | 76 | 75 (7.2) |

Subgroup phenotypes in the ADNI are from [19]. The executive functioning, language, memory, and visuospatial functioning subgroups includes the AD patients with an isolated substantial relative impairment in one of four domains. The “No domain” subgroup includes those with no domain with a substantial relative impairment (< 0.80 standard deviation units), and the “Multiple domain” subgroup is for those with multiple domains with substantial relative impairments (> 0.80).

**Table S3.** Enrichment test scores (P-values) of network module, M1-14, in human brain cell types.

| **Module** | **Temporal Lobe [10]** | | | | |  | **Prefrontal Cortex [4, 11]** | | | | | | |
| --- | --- | --- | --- | --- | --- | --- | --- | --- | --- | --- | --- | --- | --- |
|  | **NC** | **MG** | **AC** | **OC** | **EC** |  | **NC (In)** | **NC (Ex)** | **MG** | **AC** | **OC** | **EC** | **PC** |
| M1 | 1.75X10^-75^ | 0.99 | 0.99 | 0.99 | 0.99 |  | 3.33X10^-36^ | 6.92X10^-116^ | 1.00 | 1.00 | 1.00 | 1.00 | 5.73X10-1 |
| M2 | 6.77X10^-104^ | 0.92 | 0.92 | 0.99 | 0.92 |  | 4.39X10^-19^ | 4.40X10^-188^ | 1.00 | 1.00 | 1.00 | 1.00 | 1.00 |
| M3 | 9.52X10^-47^ | 0.55 | 0.98 | 0.98 | 0.98 |  | 3.55X10^-9^ | 2.57X10^-48^ | 0.30 | 1.00 | 1.00 | 1.00 | 0.44 |
| M4 | 7.13X10^-64^ | 0.99 | 0.99 | 0.99 | 0.99 |  | 1.87X10^-21^ | 5.20X10^-148^ | 1.00 | 1.00 | 1.00 | 1.00 | 0.36 |
| M5 | 0.99 | 3.55X10^-4^ | 7.05X10^-13^ | 9.04X10^-4^ | 0.03 |  | 0.03 | 1.00 | 0.03 | 1.25X10^-3^ | 2.93X10^-3^ | 1.00 | 0.77 |
| M6 | 0.90 | 1.28X10^-6^ | 3.03X10^-32^ | 0.90 | 4.24X10^-4^ |  | 0.31 | 1.00 | 2.02X10^-3^ | 1.20X10^-28^ | 1.98X10^-4^ | 1.47X10^-10^ | 9.54X10^-4^ |
| M7 | 0.99 | 0.93 | 5.15X10^-76^ | 0.99 | 0.07 |  | 0.35 | 1.00 | 0.79 | 1.32X10^-97^ | 0.79 | 1.65X10^-12^ | 1.05X10^-11^ |
| M8 | 0.83 | 0.23 | 6.00X10^-6^ | 0.06 | 6.72X10^-6^ |  | 0.79 | 1.00 | 0.08 | 2.45X10^-11^ | 0.04 | 2.75X10^-4^ | 0.16 |
| M9 | 1.00 | 0.15 | 1.68X10^-1^ | 0.10 | 4.10X10^-9^ |  | 0.16 | 0.98 | 5.31X10^-8^ | 6.03X10^-7^ | 1.36X10^-6^ | 5.09X10^-8^ | 1.73X10^-5^ |
| M10 | 1.00 | 1.65X10^-4^ | 1.00 | 1.00 | 9.93X10^-87^ |  | 1.00 | 1.00 | 4.27X10^-4^ | 0.61 | 1.00 | 2.17X10^-84^ | 1.64X10^-17^ |
| M11 | 1.00 | 1.72X10^-129^ | 1.00 | 1.00 | 1.00 |  | 1.00 | 1.00 | 1.59X10^-33^ | 1.00 | 1.00 | 0.50 | 0.88 |
| M12 | 0.30 | 0.20 | 0.99 | 0.93 | 0.99 |  | 0.29 | 1.96X10^-5^ | 0.15 | 1.00 | 1.00 | 1.00 | 0.75 |
| M13 | 0.99 | 0.96 | 0.99 | 0.96 | 0.96 |  | 0.26 | 1.00 | 4.43X10^-9^ | 0.12 | 3.15X10^-5^ | 0.03 | 0.15 |
| M14 | 1.00 | 0.45 | 1.00 | 0.84 | 0.84 |  | 0.34 | 1.00 | 2.20X10^-3^ | 0.43 | 0.37 | 1.00 | 0.89 |

NC: neurons. MG: microglia. AC: astrocytes. OC: oligodendrocytes. EC: endothelial cells. PC: oligodendrocyte progenitor cell. In: inhibitory neurons. Ex: excitatory neuron

**Table S4.** Means with standard deviation (SD) of module-based PRSs of M1-14 in Alzheimer’s disease patients.

|  | Mean | SD |
| --- | --- | --- |
| M1 | 23.3 | 0.8 |
| M2 | 1.95 | 0.2 |
| M3 | 41.2 | 3.1 |
| M4 | 7.3 | 0.56 |
| M5 | 5.11 | 0.55 |
| M6 | 0.787 | 0.25 |
| M7 | 12.8 | 0.78 |
| M8 | 12 | 0.65 |
| M9 | 6.75 | 0.54 |
| M10 | 1.77 | 0.14 |
| M11 | 13.4 | 2.7 |
| M12 | 33.4 | 1.8 |
| M13 | 4.07 | 0.4 |
| M14 | 4.67 | 0.4 |

**Table S5.** Significant associations at P<0.05 between mbPRSs and domain-specific cognitive tests in ADNI

| **mbPRS** | **Outcome** | **BETA** | **SE** | **P-value** |
| --- | --- | --- | --- | --- |
| M3 | BNTTOTAL | -0.073 | 0.010 | 1.74E-13 |
|  | CATANIMSC | -0.058 | 0.010 | 5.13E-09 |
|  | LDELTOTAL | -0.081 | 0.010 | 2.69E-16 |
|  | LIMMTOTAL | -0.083 | 0.010 | 5.16E-17 |
|  | TRAASCOR | 0.035 | 0.010 | 4.49E-04 |
|  | TRABSCOR | 0.056 | 0.010 | 2.21E-08 |
| M6 | BNTTOTAL | 0.255 | 0.117 | 2.89E-02 |
|  | CATANIMSC | 0.286 | 0.116 | 1.40E-02 |
|  | TRABSCOR | -0.257 | 0.117 | 2.80E-02 |
| M7 | BNTTOTAL | -0.083 | 0.037 | 2.79E-02 |
|  | LDELTOTAL | -0.084 | 0.037 | 2.58E-02 |
|  | TRABSCOR | 0.102 | 0.037 | 6.46E-03 |
| M9 | BNTTOTAL | -0.167 | 0.055 | 2.57E-03 |
|  | COPYSCORE | -0.097 | 0.058 | 4.91E-02 |
|  | LDELTOTAL | -0.180 | 0.055 | 1.14E-03 |
|  | LIMMTOTAL | -0.149 | 0.055 | 7.18E-03 |
|  | TRAASCOR | 0.181 | 0.055 | 1.08E-03 |
|  | TRABSCOR | 0.176 | 0.056 | 1.58E-03 |
| M10 | BNTTOTAL | -0.421 | 0.176 | 1.72E-02 |
|  | TRABSCOR | 0.369 | 0.176 | 3.67E-02 |
| M11 | BNTTOTAL | -0.087 | 0.011 | 4.87E-14 |
|  | CATANIMSC | -0.069 | 0.011 | 2.49E-09 |
|  | COPYSCORE | -0.026 | 0.012 | 2.85E-02 |
|  | LDELTOTAL | -0.102 | 0.011 | 4.93E-19 |
|  | LIMMTOTAL | -0.104 | 0.011 | 6.86E-20 |
|  | TRAASCOR | 0.055 | 0.012 | 1.79E-06 |
|  | TRABSCOR | 0.075 | 0.011 | 1.07E-10 |
| M12 | BNTTOTAL | -0.081 | 0.017 | 2.23E-06 |
|  | CATANIMSC | -0.044 | 0.017 | 1.18E-02 |
|  | LDELTOTAL | -0.070 | 0.017 | 4.55E-05 |
|  | LIMMTOTAL | -0.079 | 0.017 | 4.42E-06 |
|  | TRABSCOR | 0.034 | 0.017 | 4.70E-02 |

**Table S6.** Sample size and characteristics of cognitive tests in AD cases from the FHS

| Total, N | 134 |
| --- | --- |
| Females (%) | 86 (64.2%) |
| Age at Exam, Mean (SD) | 84.0 (8.0) |
| Cognitive Tests, Mean (SD) |  |
| TRAA | 1.9 (1.8) |
| TRAB | 6.1 (3.6) |
| VRI | 3.1 (2.3) |
| VRD | 1.6 (2.0) |
| BNT30 | 18.0 (6.9) |
| BNT30cue | 21.0 (7.4) |
| LMI | 4.2 (4.0) |
| LMD | 2.9 (3.9) |

Trail making test A/B (TRAA and TRAB); visual reproductions Immediate and Delayed Recalls (VRI and VRD), Boston naming test without cue (BNT30) and with cue (BNT30cue), and logical memory test, immediate and delayed recalls (LMI and LMD); SD: standard deviation

**Table S7.** Summary statistics of associations between mbPRSs for (M6, M9, and M12) and domain-specific cognitive tests in FHS.

| **Cognitive Domain** | **Cognitive Test** |  | **M6** | | |  | **M9** | | |  | **M12** | | |
| --- | --- | --- | --- | --- | --- | --- | --- | --- | --- | --- | --- | --- | --- |
|  |  |  | **BETA** | **SE** | **P-value** |  | **BETA** | **SE** | **P-value** |  | **BETA** | **SE** | **P-value** |
| Executive functioning | TRAA |  | 2.13 | 1.88 | 0.27 |  | 0.50 | 0.72 | 0.50 |  | -0.34 | 0.62 | 0.59 |
|  | TRAB |  | 3.16 | 3.77 | 0.41 |  | 1.93 | 1.47 | 0.20 |  | 1.38 | 1.26 | 0.29 |
| Visuospatial functioning | VRI |  | 3.84 | 1.89 | 0.05 |  | 0.11 | 0.76 | 0.88 |  | -0.42 | 0.66 | 0.52 |
|  | VRD |  | 3.68 | 1.64 | 0.03 |  | 0.47 | 0.63 | 0.47 |  | -0.04 | 0.57 | 0.94 |
| Language | BNT30 |  | 12.71 | 5.72 | 0.03 |  | 2.19 | 2.28 | 0.34 |  | -1.39 | 1.93 | 0.48 |
|  | BNT30cue |  | 14.24 | 6.12 | 0.03 |  | 2.74 | 2.44 | 0.27 |  | -1.86 | 2.07 | 0.38 |
| Memory | LMD |  | 3.05 | 3.01 | 0.32 |  | 0.12 | 1.27 | 0.92 |  | -1.52 | 1.06 | 0.16 |
|  | LMI |  | 5.54 | 3.15 | 0.09 |  | 1.18 | 1.20 | 0.33 |  | -1.16 | 1.03 | 0.27 |

Trail making test A/B (TRAA and TRAB); visual reproductions Immediate and Delayed Recalls (VRI and VRD), Boston naming test without cue (BNT30) and with cue (BNT30cue), and logical memory test, immediate and delayed recalls (LMI and LMD)

**Table S8.** Summary of gene functions in Modules, M6 and M9

| **Module** | **Gene Symbol** | **Gene Name** | **Gene Function Summary**  **Reports related with Neurological Disorder (PMID)** |
| --- | --- | --- | --- |
| M6 | ADCY2 | adenylate cyclase 2 | - Adenylate cyclase-activating G protein-coupled receptor signaling pathway (10802651)  - AD related changes in hippocampal gene expression (22982105; 22209813) |
|  | ARHGAP31 | Rho GTPase activating protein 31 | - Regulation of small GTPase mediated signal transduction  - Association with Late-Onset Alzheimer’s Disease in Gene–Gene interactions study in the Russian population (34681041) |
|  | AUTS2 | activator of transcription and developmental regulator | - Actin cytoskeleton reorganization axon extension; dendrite extension; neuron migration - Associations with intellectual disability in gene expression study (26333717)  - Identified in the GWAS for Autism spectrum disorder (17211639) |
|  | CARD8 | caspase recruitment domain family member 8 | - Activation of cysteine-type endopeptidase activity involved in apoptotic process  - Deficiency of CARD8 is associated with increased Alzheimer's disease risk in women (18841008) |
|  | CLDN10 | claudin 10 | - Bicellular tight junction assembly (25079797) |
|  | DOCK1 | dedicator of cytokinesis 1 | - Fc-gamma receptor signaling pathway involved in phagocytosis  - Missense mutations in DOCK1 were observed in autism spectrum disorder (25363768) |
|  | GNG12 | G protein subunit gamma 12 | - Cerebral cortex development (30820025) |
|  | MAPRE1 | microtubule associated protein RP/EB family member 1 | - Neurite outgrowth, axon formation, and dendritic branching (21248129; 24954409) |
|  | PCDHGB2 | protocadherin gamma subfamily B, 2 | - Homophilic cell adhesion via plasma membrane adhesion molecules (31784692) |
|  | PCDHGB4 | protocadherin gamma subfamily B, 4 | - Calcium-dependent cell-cell adhesion via plasma membrane cell adhesion molecules (31784692) |
|  | PDLIM3 | PDZ and LIM domain 3 | - Actin cytoskeleton organization  - Genetic modifiers of age at onset in early and late onset AD linkage study (26214276) |
|  | PTPRG | protein tyrosine phosphatase receptor type G | - Brain development; negative regulation of neuron projection development (26830138) |
|  | SLC6A11 | solute carrier family 6 member 11 | - Brain development; sodium ion transmembrane transport  - Relevant to Epilepsy (21752877) |
|  | SLCO1C1 | solute carrier organic anion transporter family member 1C1 | - Sodium-independent organic anion transport; transport across blood-brain barrier (26687838) |
|  | SNTB1 | syntrophin beta 1 | - Altered gene expression level in diabetes patients with AD (23595620) |
|  | ZNF423 | zinc finger protein 423 | - Notch signaling pathway; cerebellar granule cell precursor proliferation  - Identified in gene-based analysis for AD (31283791) |
| M9 | CBFB | core-binding factor subunit beta | - Cell maturation; negative regulation of CD4-positive, alpha-beta T cell differentiation  - Associations with AD and Lewy body dementia in gene expression study (29503795; 33257666) |
|  | CCDC88C | coiled-coil domain containing 88C | - Apical constriction; cytoplasmic microtubule organization (30679421) |
|  | DDX60L | DExD/H-box 60 like | - Anti-viral immunity; hepatitis C virus replication |
|  | DISC1 | DISC1 scaffold protein | - Neuron migration; positive regulation of axon extension (25224257) |
|  | FRMD6 | FERM domain containing 6 | - Actomyosin structure organization  - GWAS of hippocampal atrophy and cognitive decline rates in AD patients (22190428; 24092460; 23535033) |
|  | GLIS3 | GLIS family zinc finger 3 | - Negative regulation of transcription by RNA polymerase II - Genetic association with elevated levels of cerebrospinal fluid tau and phosphorylated tau (23562540; 28911974) |
|  | ITGBL1 | integrin subunit beta like 1 | - Cell adhesion mediated by integrin  - Differential gene expression study of AD (28373841) |
|  | OAS2 | 2'-5'-oligoadenylate synthetase 2 | - Innate immune response to viral infection.  - Pleiotropy analysis of AD and ischemic stroke (30859738) |
|  | RNF213 | ring finger protein 213 | - Identified in the P301S mouse model of tauopathy (30696811) |
|  | SLC25A30 | solute carrier family 25 member 30 | - Phosphate ion transmembrane transport  - Differential gene expression study of anxiety/depression-like disorder (30537945) |

**Table S9.** Top-ranked gene ontology (GO) findings from Ingenuity Pathway Analysis.

| **Module** | **GO - Diseases or Functions Annotation** | **P-value** |
| --- | --- | --- |
| M6 | Morphology of nervous system | 4.02E-08 |
| M6 | Abnormal morphology of nervous system | 7.83E-07 |
| M6 | Morphology of neuroglia | 9.58E-07 |
| M6 | Proliferation of neuronal cells | 4.80E-06 |
| M6 | Quantity of neurons | 1.66E-05 |
| M6 | Development of central nervous system | 3.31E-05 |
| M6 | Differentiation of neuroglia | 4.74E-05 |
| M6 | Formation of brain | 7.01E-05 |
| M6 | Sensory system development | 7.72E-05 |
| M6 | Differentiation of astrocytes | 0.000083 |
| M9 | Development of vasculature | 3.83E-09 |
| M9 | Angiogenesis | 3.02E-08 |
| M9 | Vasculogenesis | 2.48E-07 |
| M9 | Abnormal morphology of cardiovascular system | 5.76E-06 |
| M9 | Morphology of cardiovascular system | 7.01E-06 |
| M9 | Cardiogenesis | 1.54E-05 |
| M9 | Morphology of heart | 6.46E-05 |
| M9 | Morphology of cardiac muscle | 7.62E-05 |
| M9 | Abnormal morphology of vasculature | 9.56E-05 |
| M9 | Abnormal morphology of heart | 1.25E-04 |

**Table S10.** Differential expression of AD associated genes in M6 and M9 between AD and control brains in FHS/BUADRC brains.

| **Module** | **Gene** | **Z-score** | **P-value** |
| --- | --- | --- | --- |
| M6 | *ADCY2* | 0.84 | 0.4 |
| M6 | *ARHGAP31* | 4.77 | 1.80E-06 |
| M6 | *AUTS2* | 0.28 | 0.78 |
| M6 | *CARD8* | 2.77 | 5.60E-03 |
| M6 | *CLDN10* | 3.36 | 7.70E-04 |
| M6 | *DOCK1* | 5.12 | 3.00E-07 |
| M6 | *GNG12* | 4.38 | 1.20E-05 |
| M6 | *MAPRE1* | 4.77 | 1.80E-06 |
| M6 | *PCDHGB2* | 1.37 | 0.17 |
| M6 | *PCDHGB4* | -0.53 | 0.60 |
| M6 | *PDLIM3* | 3.57 | 3.60E-04 |
| M6 | *PTPRG* | -0.62 | 0.53 |
| M6 | *SLC6A11* | 1.66 | 0.098 |
| M6 | *SLCO1C1* | 0.24 | 0.81 |
| M6 | *SNTB1* | 2.71 | 6.80E-03 |
| M6 | *ZNF423* | 4.07 | 4.70E-05 |
| M9 | *CBFB* | 2.94 | 3.20E-03 |
| M9 | *CCDC88C* | 2.93 | 3.40E-03 |
| M9 | *COLEC12* | 2.36 | 0.02 |
| M9 | *DDX60L* | 0.58 | 0.56 |
| M9 | *DISC1* | 2.82 | 4.90E-03 |
| M9 | *FRMD6* | 1.69 | 0.09 |
| M9 | *GLIS3* | 3.06 | 2.20E-03 |
| M9 | *ITGBL1* | 1.33 | 0.18 |
| M9 | *OAS2* | 1.15 | 0.25 |
| M9 | *RNF213* | 2.7 | 0.01 |
| M9 | *SLC25A30* | 4.52 | 6.30E-06 |

Analysis results were from [4]*.*

**Table S11.** Significant associations at P<0.05 between expression of genes in M6 and M9 modules and AD-related traits in FHS/BUADRC brains

| **Module** | **Outcome** | **Gene Expression** | **Beta** | **SE** | **P-value** |
| --- | --- | --- | --- | --- | --- |
| M6 | CERAD | *DOCK1* | -0.078 | 0.039 | 4.39E-02 |
|  | Braak Stages | *DOCK1* | 0.080 | 0.036 | 2.48E-02 |
|  |  | *PTPRG* | -0.081 | 0.029 | 5.26E-03 |
|  | Aβ42 | *CARD8* | 0.093 | 0.041 | 2.45E-02 |
|  |  | *CLDN10* | 0.156 | 0.071 | 2.85E-02 |
|  |  | *MAPRE1* | 0.091 | 0.035 | 9.20E-03 |
|  | pTau181/tTau ratio | *MAPRE1* | 0.100 | 0.037 | 8.65E-03 |
|  |  | *SLC6A11* | 0.163 | 0.077 | 3.71E-02 |
|  | pTau231/tTau ratio | *ADCY2* | 0.180 | 0.057 | 1.96E-03 |
|  |  | *ARHGAP31* | 0.118 | 0.050 | 2.03E-02 |
|  |  | *AUTS2* | 0.085 | 0.036 | 2.02E-02 |
|  |  | *DOCK1* | 0.090 | 0.041 | 2.91E-02 |
|  |  | *GNG12* | 0.106 | 0.048 | 3.03E-02 |
|  |  | *MAPRE1* | 0.080 | 0.034 | 1.93E-02 |
|  |  | *PDLIM3* | 0.124 | 0.050 | 1.46E-02 |
|  |  | *SLC6A11* | 0.139 | 0.069 | 4.53E-02 |
|  |  | *ZNF423* | 0.091 | 0.043 | 3.68E-02 |
|  | C4a | *ADCY2* | 0.221 | 0.100 | 2.83E-02 |
|  |  | *AUTS2* | 0.363 | 0.161 | 2.55E-02 |
|  |  | *CLDN10* | 0.189 | 0.080 | 1.89E-02 |
|  |  | *SLCO1C1* | 0.161 | 0.074 | 3.02E-02 |
|  | C4b | *ARHGAP31* | 0.236 | 0.120 | 4.96E-02 |
|  |  | *CARD8* | 0.298 | 0.143 | 3.87E-02 |
|  |  | *DOCK1* | 0.382 | 0.149 | 1.09E-02 |
|  | PSD95 | *PCDHGB4* | -0.131 | 0.051 | 1.18E-02 |
| M9 | Aβ_42_ | *DISC1* | 0.105 | 0.032 | 1.49E-03 |
|  | Braak Stages | *FRMD6* | 0.079 | 0.033 | 1.80E-02 |
|  |  | *ITGBL1* | 0.112 | 0.050 | 2.37E-02 |
|  |  | *SLC25A30* | 0.061 | 0.029 | 3.86E-02 |
|  | pTau231/tTau ratio | *CBFB* | 0.099 | 0.044 | 2.52E-02 |
|  |  | *GLIS3* | 0.149 | 0.058 | 1.07E-02 |
|  |  | *RNF213* | 0.139 | 0.046 | 2.87E-03 |
|  |  | *SLC25A30* | 0.080 | 0.031 | 1.03E-02 |
|  | C4a | *CCDC88C* | 0.258 | 0.130 | 4.87E-02 |
|  |  | *DISC1* | -0.573 | 0.176 | 1.38E-03 |

**Figure S1.** Correlations of 3 gwPRSs (NP, NFT, and AD-risk) and 11 mbPRSs. This correlation matrix graphically represents the pair-wise correlation coefficient (r) for all PRSs tested. According to the color scale on the right side of the matrix, positive and negative correlations are indicated in shades of blue and red, respectively. The size of the circles is relative to the correlation strength.

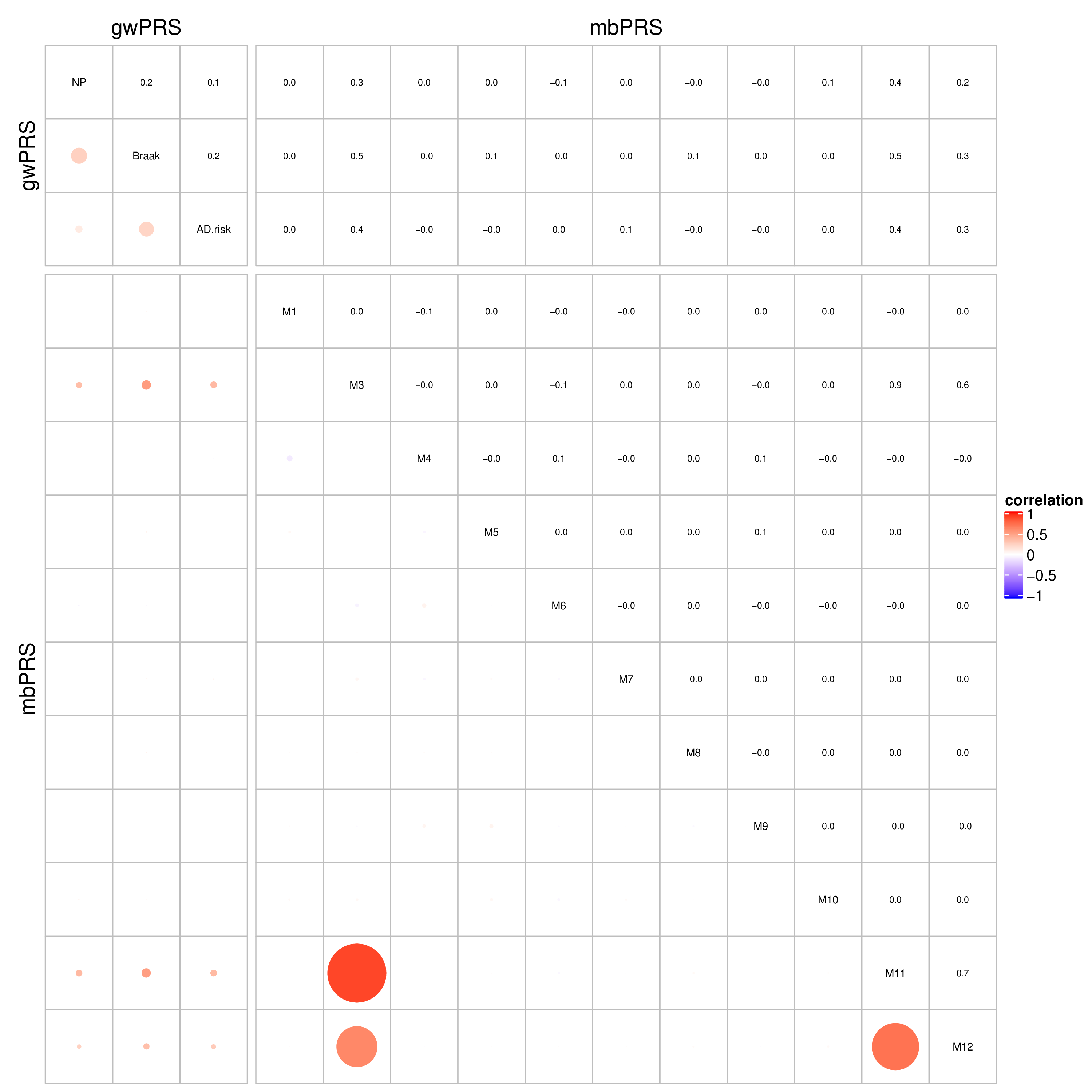
